# Priority Setting for Multicenter Research Among Adults with Cerebral Palsy: A Qualitative Study

**DOI:** 10.1101/2024.08.28.24312747

**Authors:** Cristina A. Sarmiento, Mary Gannotti, Jocelyn Cohen, Edward Hurvitz

## Abstract

**Purpose:** Identify priorities for adult cerebral palsy (CP) research by engaging individuals with lived experience, clinical investigators, and community leaders.

**Materials and Methods:** Qualitative descriptive study using iterative focus groups, followed by inductive thematic analysis. Participants included adults with CP and caregivers, clinical investigators, and community leaders in the CP and disability spaces. We explored research priorities among three research areas identified a priori– bone health, kidney health, and preventive care.

**Results:** We conducted four focus groups (20 participants with lived experience; 10 clinical investigators; 9 community leaders). Most participants felt all topic areas were very important, though preventive care emerged as the top priority. We identified three overarching themes that cut across the various research areas discussed: patient and provider knowledge gaps; a precision medicine approach for adult CP care; and the need to address ableism.

**Conclusions:** Adults with CP face unique healthcare needs and risks as they age, and the evidence base to guide their care lags significantly behind. Our study identified preventive care as the top research priority for the adult CP research agenda. Next steps in this line of research should focus on interventions to facilitate primary and preventive care interactions for adults with CP.

## Introduction

The lifespan for people with cerebral palsy (CP) is increasing, and the population of aging adults with CP is commensurately growing.^1^ Up to 90% of individuals with CP will reach adulthood, and with medical and technological advances, the greatest gains are in those with severe CP.^1^ Adults with CP have unique health needs and risks, with higher rates of chronic conditions at younger ages than the general population.^2–8^ However, adults with CP face fragmented care, difficulty finding comfortable or knowledgeable providers, and often feel like they guide their own care without sufficient resources, particularly after leaving the pediatric healthcare system.^9–11^ Prior epidemiological studies have used population health services data to establish risk for chronic disease, condition severity, and factors associated with mortality.^5,7,12–14^ Kidney disease, bone health, and primary and preventive care are all major areas of concern.^15–20^ Regarding bone health, fractures are common in both men and women with CP and lead to both increased morbidity and mortality.^21^ Improved screening and initiation of treatment for adults with CP at high risk for fracture could reduce the risk of fracture and associated sequelae. Adults with CP also have an increased risk of chronic kidney disease compared to adults without CP.^17,18^ However, early diagnosis and initiation of treatment is complicated by low muscle mass and lower baseline creatinine levels in adults with CP, which may result in delays in diagnosis.^22^ Given these risks facing adults with CP, in addition to greater risks of many other chronic illnesses, primary care and preventive screening can facilitate earlier detection and treatment of many chronic conditions. However, adults with disabilities, including CP, receive lower rates of many preventive screenings than the general population.^23–25^ However, studies from large data only identify problems; addressing them requires a different approach.

Collaboration between individuals with CP, clinical investigators, and community leaders is necessary to advance scientific and clinical knowledge to move toward improving health outcomes.^26^ Engaging individuals with lived experience in all stages of the research process facilitates the production of knowledge that is relevant and important to those with lived experience, including individuals with lifespan disabilities.^27,28^ The Cerebral Palsy Research Network (CPRN) is a learning health network for CP consisting of more than 30 clinical sites and bringing together clinicians, researchers, and persons with lived experience (i.e., individuals with CP, family members, caregivers) to improve health outcomes for people with CP.^29^ Leveraging the CPRN infrastructure can facilitate both the co-creation of an adult CP research agenda and the development of a multicenter research program focused on adults and aging with CP. With the overall goal of addressing negative health outcomes in adults with CP, the objective of this study was to identify priorities for the adult CP research agenda by engaging individuals with lived experience, clinical investigators, and community leaders.

## Materials and Methods

### Study Design

This qualitative descriptive study used iterative focus groups to gather input from people with lived experience, community organizations, clinicians, and researchers regarding priorities for this research agenda. The Colorado Multiple Institutional Review Board determined our study to be exempt from review (Protocol #23-1653). We report our qualitative methods and findings in accordance with the Standards for Reporting Qualitative Research.^30^

### Participants and Procedures

We conducted three rounds of focus groups that included knowledge sharing and education related to potential research areas, followed by participant input on recruitment, prioritization, and implementation. The first round of focus groups included adults with CP and caregivers of adults with CP; the second round of focus groups included clinical investigators; and the third round of focus groups included community leaders, with details of focus group composition and eligibility criteria detailed in table 1. We used purposeful sampling,^31–33^ including maximum variation and snowball sampling approaches for diversity of participant background based on characteristics in table 1. In maximum variation sampling, key variables are selected by the research team and then cases that vary from each other as much as possible are identified, allowing us to identify meaningful patterns from a diverse sample.^33^ In snowball sampling, the study team seeks information from key informants about other information-rich cases or participants.^33^

**Table 1.** Focus group participants, recruitment, and eligibility criteria.

| Focus Group Participants | Recruited from | Eligibility criteria | Purposefully sampled for diversity of | Number and size of focus groups |
| --- | --- | --- | --- | --- |
| Adults with CP and caregivers | Study team clinical sites<br>CPRN, CPARF stakeholder groups | 30 years and older* + diagnosis of CP<br>-OR- Caregiver of adult with CP | Gender<br>Race and ethnicity<br>Functional mobility level^ | 2 focus groups<br>8-10 participants each |
| Clinical investigators | CPRN<br>Other CP-focused clinical sites | Self-identified clinician and/or investigators who regularly treats adults with CP or is involved in transition from pediatric to adult care<br>Interest in multicenter research program | Clinical role (e.g., MD, PT)<br>Specialty (e.g., neurology, PM&R) | 1 focus group<br>8-10 participants |
| Community leaders and advocates | CP and disability community organizations<br>Social media advocates in CP and disability spaces | Potential to / interest in using platform to promote adult CP research | Organizations/ platforms | 1 focus group<br>8-10 participants |

\*We used a minimum age of 30 years for adults with CP as we were seeking participant perspectives on adult health issues and adult healthcare and therefore sought participants with several years of experience in adult healthcare settings.
^based on Gross Motor Function Classification System level
CPARF – Cerebral Palsy Alliance Research Foundation
MD – Medical doctor
PM&R – Physical medicine & rehabilitation
PT – Physical therapist

The study team developed the focus group guides based on the study objectives, prior research, and input from CPRN clinicians and community members during webinars detailing this study’s proposed methods. Focus groups took place over Zoom videoconferencing, lasted approximately 90 minutes, and were facilitated by a non-clinician member of the study team who is an adult with CP (JC) to minimize the potential bias of having a clinician facilitator. Verbal consent was obtained at the start of each focus group. Within each group, the study team first shared knowledge about three potential research areas related to adults with CP (bone health and fracture risk, renal function impairment, and primary care and preventive care gaps). These areas were selected by the research team as high need, high potential impact areas for the reasons described above, as well the ability to leverage CPRN’s multisite network for future interventional studies in these areas. Then, the facilitator guided conversation to glean participant perspectives on prioritization of these research areas, potential barriers and facilitators to each area’s implementation, and recruitment strategies. In the second and third rounds of focus groups, results from the prior focus groups were summarized and shared to iteratively build upon prior discussions. During focus groups with participants with lived experience and clinical investigators, we also assessed perceived importance and ranked prioritization of these research areas via Zoom polling function. Focus groups were audio-recorded with permission from participants and professionally transcribed (Landmark Associates; Phoenix, AZ). Transcripts were de-identified prior to analysis. Participants also entered self-reported demographic data into a secure REDCap electronic database hosted at the University of Colorado Anschutz ahead of the focus groups.^34^

### Data Analysis

We used a qualitative descriptive approach that allowed us to explore, understand, and describe the varied perspectives of participants.^35–37^ The study team (CS, MG, EH) coded transcripts inductively, creating codes based on emergent data.^38^ The team met regularly to reconcile and calibrate coded transcripts until a final codebook was developed.^39^ All transcripts were double coded by at least two of the three coding team members. We then categorized and grouped together all the responses and dialogue of the participants in the focus group. Using traditional thematic analysis, we reviewed and summarized quotations and identified salient themes.^39,40^

## Results

We conducted four focus groups (20 participants with lived experience; 10 clinical investigators; 9 community leaders). Demographic characteristics of participants are listed in tables 2-4. Participants with lived experience had diverse levels of mobility function, clinical investigators represented a variety of specialties, and community organization leaders served both CP-specific and broader disability communities across the country.

**Table 2.**
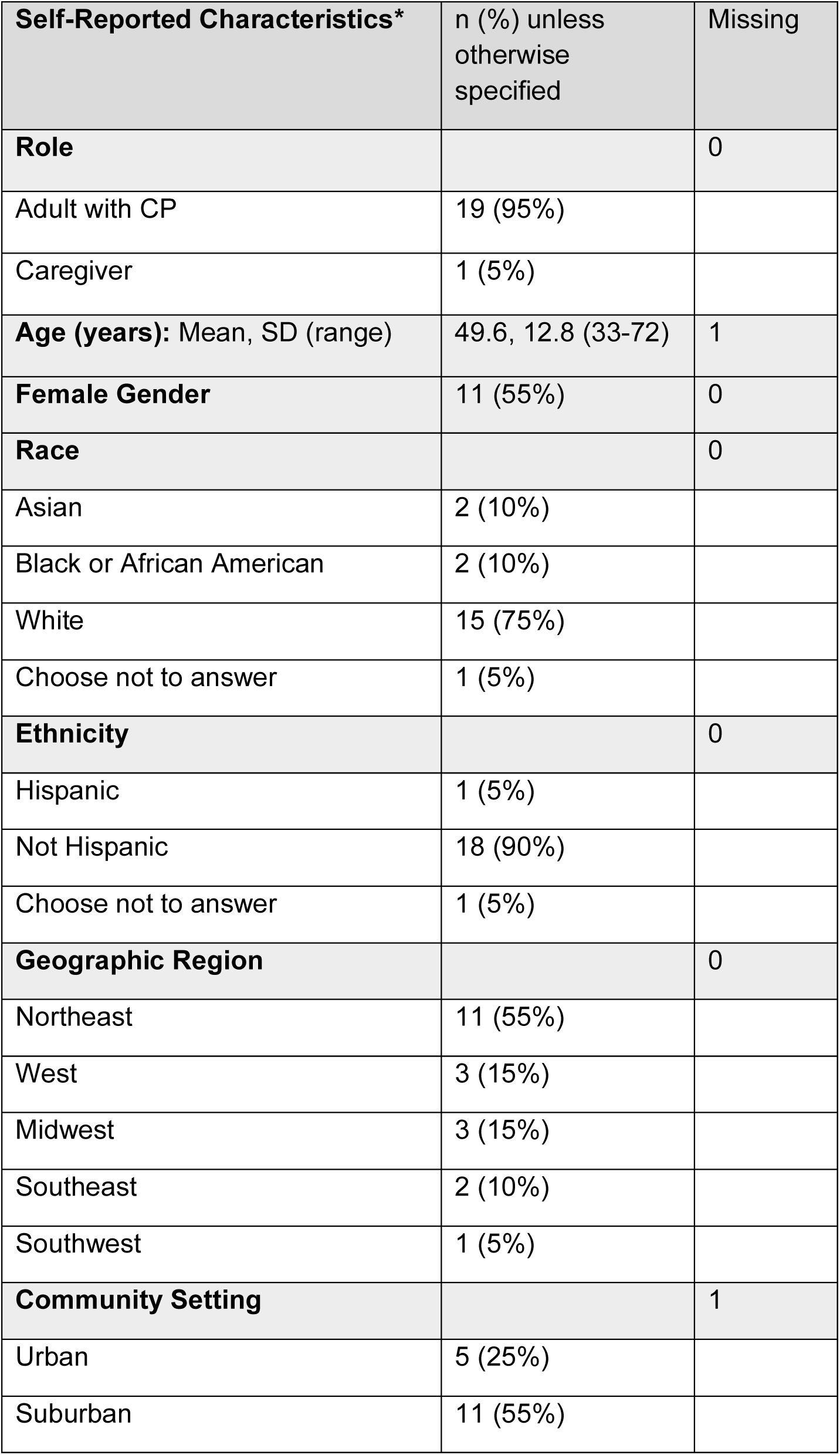
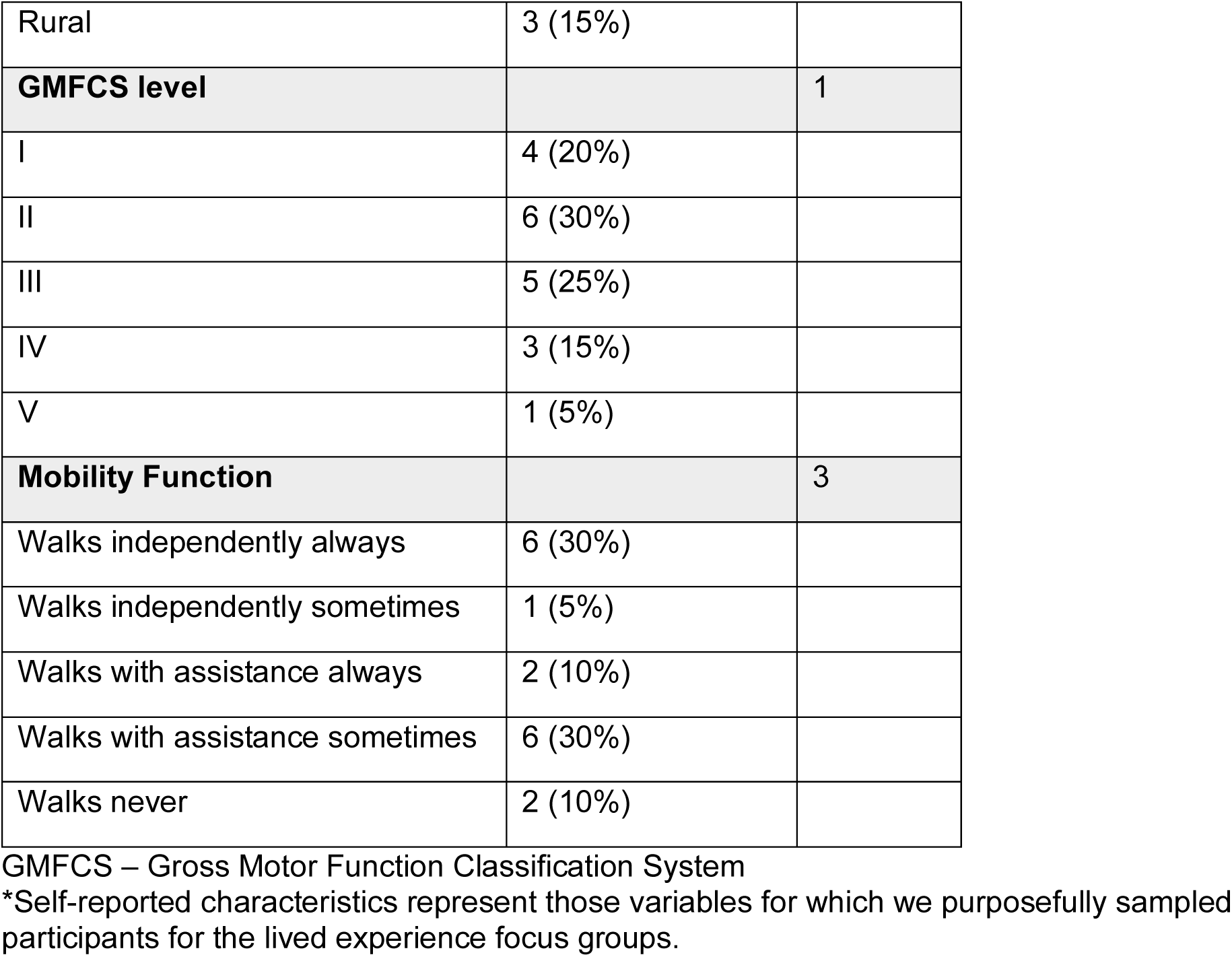
Characteristics of adults with cerebral palsy and caregivers participating in focus groups (N=20).

| **Self-Reported Characteristics\*** | n (%) unless otherwise specified | Missing |
| --- | --- | --- |
| **Role** |  | 0 |
| Adult with CP | 19 (95%) |  |
| Caregiver | 1 (5%) |  |
| **Age (years):** Mean, SD (range) | 49.6, 12.8 (33-72) | 1 |
| **Female Gender** | 11 (55%) | 0 |
| **Race** |  | 0 |
| Asian | 2 (10%) |  |
| Black or African American | 2 (10%) |  |
| White | 15 (75%) |  |
| Choose not to answer | 1 (5%) |  |
| **Ethnicity** |  | 0 |
| Hispanic | 1 (5%) |  |
| Not Hispanic | 18 (90%) |  |
| Choose not to answer | 1 (5%) |  |
| **Geographic Region** |  | 0 |
| Northeast | 11 (55%) |  |
| West | 3 (15%) |  |
| Midwest | 3 (15%) |  |
| Southeast | 2 (10%) |  |
| Southwest | 1 (5%) |  |
| **Community Setting** |  | 1 |
| Urban | 5 (25%) |  |
| Suburban | 11 (55%) |  |
| Rural | 3 (15%) |  |
| **GMFCS level** |  | **1** |
| I | 4 (20%) |  |
| II | 6 (30%) |  |
| III | 5 (25%) |  |
| IV | 3 (15%) |  |
| V | 1 (5%) |  |
| **Mobility Function** |  | **3** |
| Walks independently always | 6 (30%) |  |
| Walks independently sometimes | 1 (5%) |  |
| Walks with assistance always | 2 (10%) |  |
| Walks with assistance sometimes | 6 (30%) |  |
| Walks never | 2 (10%) |  |

GMFCS – Gross Motor Function Classification System
\*Self-reported characteristics represent those variables for which we purposefully sampled participants for the lived experience focus groups.

**Table 3.**
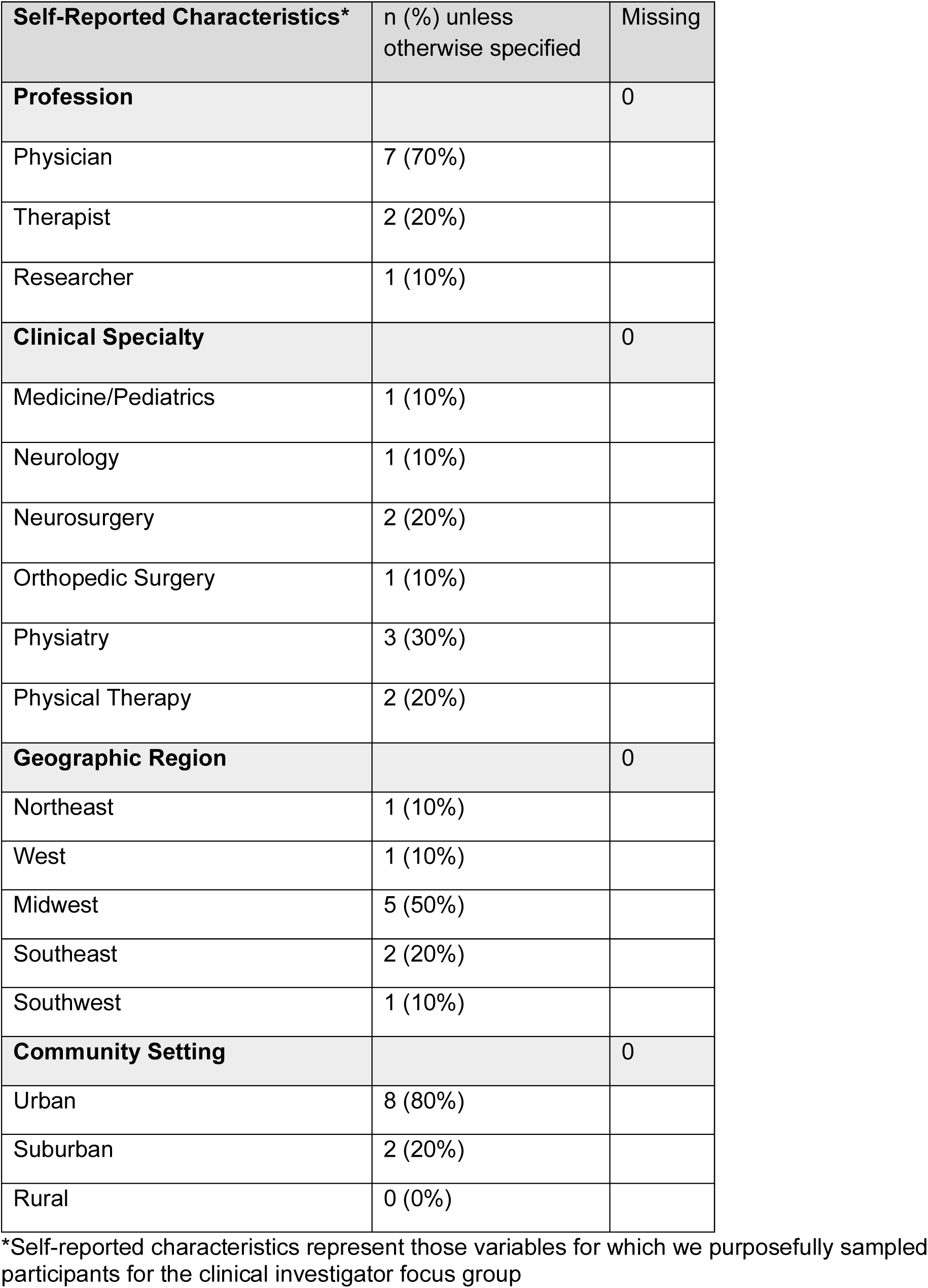
Characteristics of clinical investigators participating in focus groups (N=10).

**Table 4.** Characteristics of community leaders participating in focus groups (N=9).

| **Self-Reported Characteristics\*** | n (%) unless otherwise specified | Missing |
| --- | --- | --- |
| **Community Organization Focus** |  | 1 |
| Cerebral palsy | 5 (56%) |  |
| General disability | 2 (22%) |  |
| Intellectual and developmental disability | 1 (11%) |  |
| **Number of Different Organizations Represented^** | 6 | 1 |
| **Geographic Region** |  | 1 |
| Northeast | 4 (44%) |  |
| West | 0 (0%) |  |
| Midwest | 1 (11%) |  |
| Southeast | 1 (11%) |  |
| Southwest | 2 (22%) |  |
| **Community Setting** |  | 2 |
| Urban | 3 (33%) |  |
| Suburban | 4 (44%) |  |
| Rural | 0 (0%) |  |

\*Self-reported characteristics represent those variables for which we purposefully sampled participants for the community leader focus group.
^Remainder of participants were unaffiliated people with cerebral palsy (e.g., social media influencer)

Those who participated in the lived experience and clinical investigator focus groups were asked to rate the importance of each of the three discussed research areas (bone health, kidney health, preventive care) on a scale from 1-5 (1 = not important; 5 = extremely important) and then to rank these areas from highest to lowest priority for the adult CP research agenda. Overall, all three topic areas were felt to be very important to most participants who voted (figure 1). However, most participants ranked preventive care as the highest priority (n=15/29, 52%; figure 2) with bone health also ranked highly (n=11/29 ranked bone health highest, 38%).

**Figure 1.**
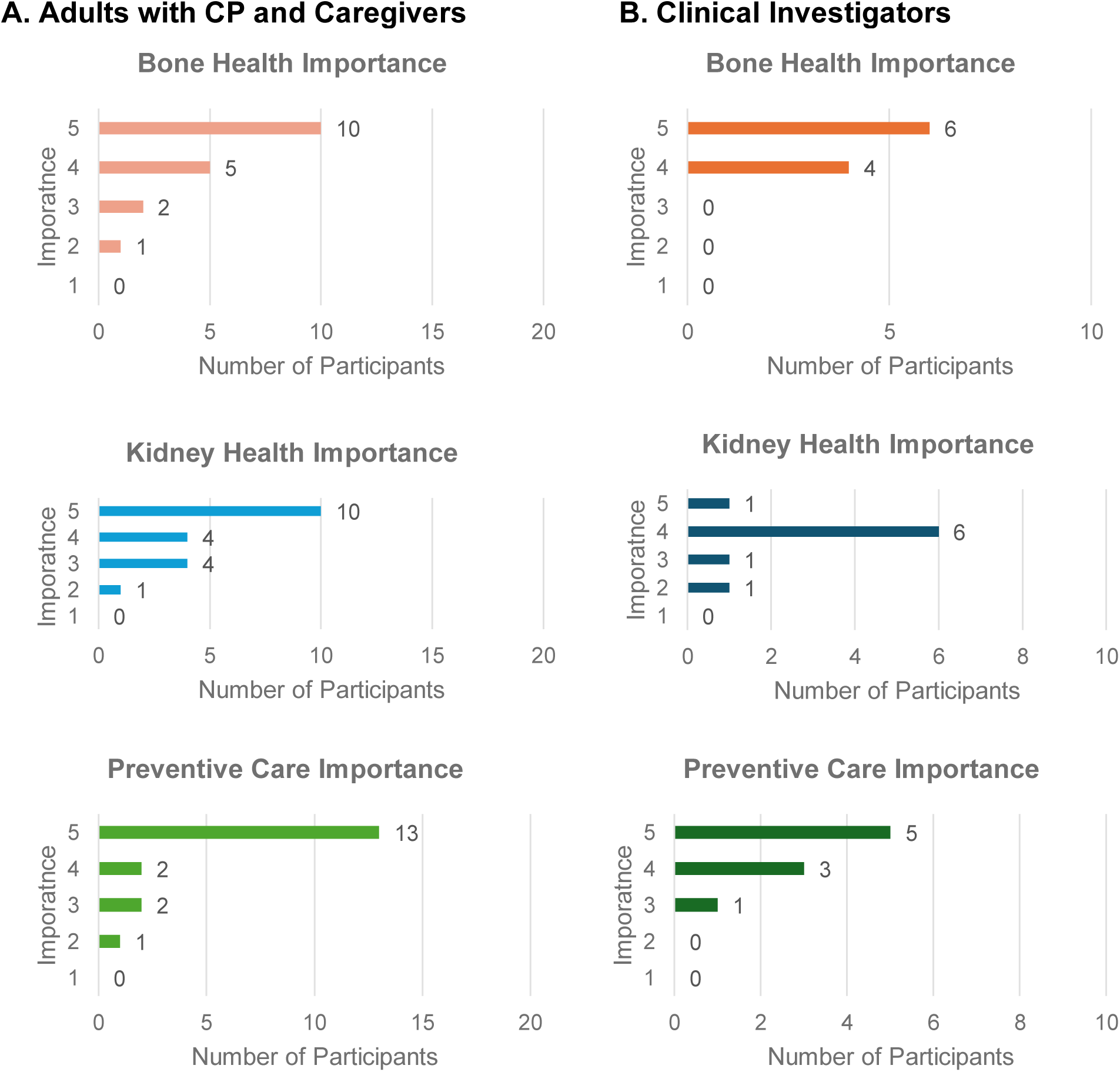
Participants with lived experience (1A) and clinical investigators (1B) rate the importance of bone health, kidney health, and preventive care for the adult cerebral palsy research agenda (1 = not important; 5 = extremely important). X-axis shows number of participants out of the total number of participants (N=20 participants with lived experience; N=10 clinical investigators). One clinical investigator participant had to leave the focus group early and thus did not vote in the kidney health or preventive care polls.

**Figure 2.**
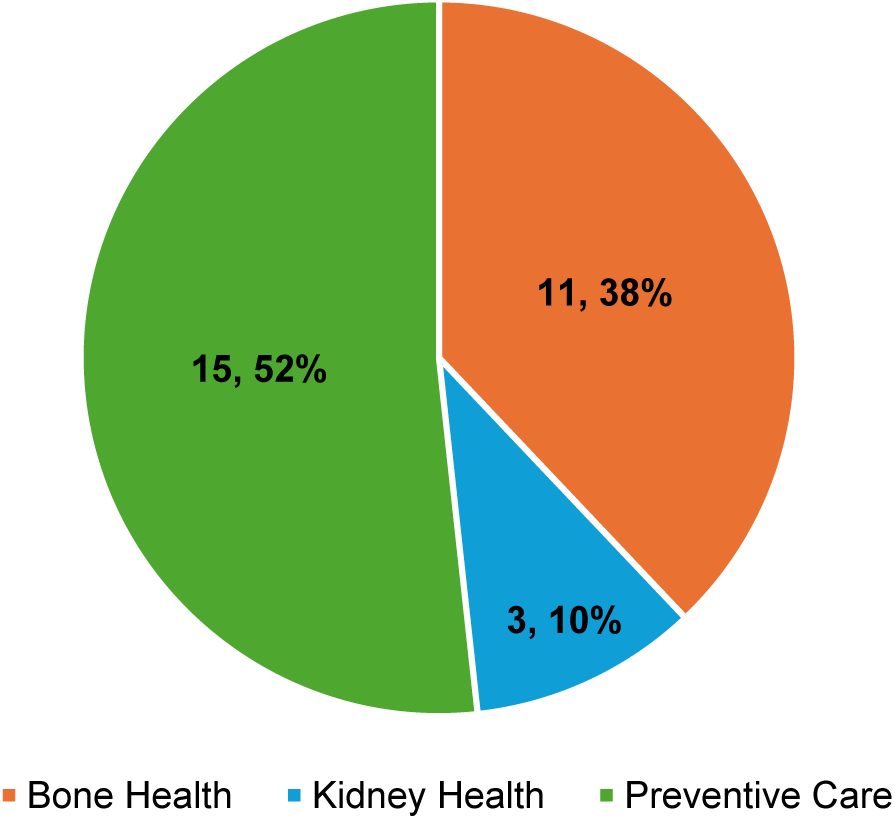
Highest priority research topic according to participants with lived experience and clinical investigators (N=29; one clinical investigator participant had to leave the focus group early and thus did not participate in this poll). Presented as n(%).

From our thematic analysis, we identified three overarching themes that cut across the various research areas discussed: patient and provider knowledge gaps; a precision medicine approach for adult CP care; and the need to address ableism. Table 5 presents each theme with illustrative quotes.

**Table 5.** Identified themes and illustrative quotes, organized by research topic area.

|  | **Theme 1:**<br>**Patient and Provider**<br>**Knowledge Gaps** | **Theme 2:**<br>**A Precision Medicine**<br>**Approach for Adult**<br>**CP Care** | **Theme 3:**<br>**The Need to Address**<br>**Ableism** |
| --- | --- | --- | --- |
| **Bone Health** | A year ago, when I had a very mild fall but ended up fracturing my arm, and it was the second fracture I had in less than a year. It's very frustrating because it all comes down to awareness. (Participant 3, adult with CP)<br>I never even heard of a DEXA scan. I have no idea what it is. I never heard of it. No one ever mentioned it to me. (Participant 8, adult with CP) | I think that as we think about looking at this issue in the future, we make sure we fold fall history, fear of falling, avoidance of behaviors due to prior falls. (Participant 25, clinical investigator) | I have [diffuse] osteopenia, and nobody wants to address it, as well as a few other things that nobody wants to address either. You're sort of left here knowing that your health's going down the tank, and there's not much you can do about it. That's my frustration. (Participant 2, adult with CP) |
| **Kidney Health** | I'm a nurse, and I didn't even realize that that affected the creatinine level and how much muscle mass we have or don't have. I guess my point is that, even as a medical professional, that didn't even cross my mind. I did not even correlate that relation between the muscle mass and the creatinine levels. There's definitely a huge barrier right there, just in that aspect, for me. (Participant 4, adult with CP) | I would also add, it's not just screening tools, but what is an appropriate baseline [creatinine], and what would be an appropriate amount of increase and/or decrease...It could also be different for different GMFCS levels. We don't necessarily know that yet. I assume. I think that would be an interesting question to look into. (Participant 12, adult with CP) | I completely agree about the under hydration...They're only gonna change the diaper twice a day and so they keep them dehydrated...That decade after decade of dehydration probably contributes. (Participant 21, clinical investigator) |
| **Preventive Care** | My primary care doctor is more or less of a joke when it comes to my CP. (Participant 8, adult with CP)<br>When we see our primary care provider, it's only because he needs a vaccine, or he has an infection or whatnot. I'm not seeing any preventative care... (Participant 11, caregiver)<br>My experience is that I spent an awful lot of time, in my lifetime, educating and advocating primary care, and | I think the preventive care's the biggest issue, trying to get the providers and the insurance company to realize that the general standard does not apply to us always. They need to think outside of the box. That's pretty much like pullin' teeth 'cause, at least from mine, they just blow you off. It's irritating. (Participant 7, adult with CP) | With all due respect, the residents and people who are coming up through the next generation of physicians, they are barely aware of CP or disability in general. (Participant 1, adult with CP)<br>One thing in particular that I think is often dismissed when I have a concern about it is hypertension and high resting heart rate in adults with CP, which to me is indicating that the cardiovascular system is working harder than it |
|  | other doctors that I see, about CP... I guess my experience has been that I've had to be my own best advocate. (Participant 1, adult with CP)<br>I'm always in my line of work, always supporting our consumer community in learning how to advocate for themselves as it relates to being able to describe their disability and what they may be experiencing that's different from their disability, the regular experiences that they deal with as related to their disability. (Participant 38, community leader) |  | should be. I'm often told, "Well, we just expect that in this population," and nothing is done or considered about that. (Participant 24, clinical investigator) |

DEXA – dual-energy X-ray absorptiometry GMFCS – Gross Motor Function Classification System

### Theme 1: Patient and Provider Knowledge Gaps

Participants across focus groups discussed provider’s simultaneous lack of awareness of and discomfort with the unique risks adults with CP face. Many participants with lived experience also shared during their focus group that they themselves were learning about these risks for the first time. Participants emphasized the need for education to increase provider awareness, but also to educate adults with CP directly to empower them to be advocates in their care. One participant clearly articulated the challenge as an adult with CP, since CP is often viewed as a childhood disability. This participant, along with several others, highlighted the need to educate individuals with CP about these risks:

> I think most people feel like [CP is] just a childhood thing. It’s not. It’s a lifelong condition, and the lack of providers in the adult world to take care of us and to really know and understand what we’re going through on a day-to-day [basis] is really, really frustrating… All these other complex things that we just talked about, like the bone health and the kidney stuff and all the preventative screening, that we’re at a higher risk for all of those because of our CP, nobody told us any of that. How are we supposed to know to advocate for ourselves, not just as an adult getting older in age but as an adult with CP who are now at a higher risk for all of these other things? – Participant 4, adult with CP

Several participants with lived experience as well as clinical investigators expressed that improving medical and allied health professional training could be a way to not only combat knowledge gaps, but also improved provider comfort in treating adults with CP and other disabilities. As one participant shared:

> This is so incredibly important, this whole preventive medicine, preventive care for adults with disabilities and particularly that piece about individuals not being able to find a primary care physician that really understands cerebral palsy and is willing to take time with the individual to discuss what their issues are. That leads back into the training. -- Participant 32, community leader

A clinical investigator participant agreed, sharing:

> The way we set up our trainings out of primary care doctors for adults, they never see childhood or rarely see childhood disabilities and so it’s a huge problem. -- Participant 22, clinical investigator

In addition to training for medical students, training for physicians in residency and fellowship was another identified area in need of significant improvement by clinicians. One participant reflected on how separated pediatric and adult training for many specialties is, and how this contributes to lack of provider knowledge and disparities in care for adults with lifelong disabilities. Participants cited increased funding for and changes to the requirements of residency training to incorporate lifelong disability care as potential opportunities to improve residency training.

### Theme 2: A Precision Medicine Approach for Adult CP Care

Participants expressed the need for a “precision medicine” approach to adult CP care, or CP-specific care guidelines that take into account an individual’s risk factors (functional level, age, sex, medical co-morbidities, etc.) to guide prevention, screening, and treatment decisions. One participant wondered how mobility function (based on Gross Motor Function Classification System (GMFCS) levels) affected bone density and the need for screening:

> I was just wondering, have you guys talked about considering the GMFCS levels? I’m a pretty high functioning level one, and so I’m up and moving a lot. Obviously, that’s way different than a level four or five. Is that somethin’ that you guys are considering when you’re talking about who to scan and who to think about this? Obviously, that’s gonna affect—my bone density’s gonna be a little bit different than somebody who’s a three or a four or a five. I was just curious about that.’ -- Participant 4, adult with CP

Examples of a precision medicine approach came out across all research topics. For instance, regarding bone density screening, how can an individual’s age, sex, mobility status, medication history, fall risk, etc., be factored in to determine how often they should undergo screening? Regarding kidney disease, how does one’s medication use and medical history (including bowel and bladder issues) affect their risk of renal disease, and how does their muscle mass and mobility status impact interpretation of markers of renal disease like creatinine? All of these questions, and more, affect primary care and preventive screening. As such, a call for comprehensive, CP-specific care guidelines was made by participants across focus groups. However, in lieu of CP-specific guidelines, participants expressed that at a minimum, adults with CP should consistently receive the screenings and management recommended for the general population.

### Theme 3: The Need to Address Ableism

Unfortunately, several participants also noted ableism as a barrier to care. One participant elaborated:

> A lotta times, doctors don’t wanna even deal — this is a horrible, horrible reality, but a lotta times, doctors don’t wanna deal with people with CP because they don’t understand the way that we move. They don’t understand us when we talk. -- Participant 1, adult with CP

A couple of participants with lived experience were concerned that their communication impairments, such as dysarthria, impacted provider-patient relationships, and one was concerned their speech may cause a provider to assume they are “drunk” (Participant 19, adult with CP). Several community leaders shared that barriers to care include ableist assumptions, like people with disabilities are not sexually active or won’t have children.

> Then there’s some practitioners that put up barriers saying that well, this person doesn’t need this [preventive] care because they’re not going to child bear or they’re not—they don’t have sex, those kinds of things. I think part of it is dispelling those—a lot of it is dispelling those ableist attitudes. -- Participant 38, community leader

Several clinical investigators also highlighted ableism as an urgent issue, citing enhanced medical and health profession training as a potential way to mitigate this for future generations of clinicians, as detailed above.

While these three themes emerged across focus groups and topic areas, they were informed by participant experiences and perspectives that arose during discussion of the following three potential research topics. Participant perspectives related to each of these topic areas are discussed below, with participant-identified research questions related to these topics listed in table 6.

**Table 6.** Participant-identified research questions, organized by research topic area.

|  | **Risk Assessment/**<br>**Epidemiology** | **Prevention/**<br>**Education** | **Screening** | **Treatment/**<br>**Management** |
| --- | --- | --- | --- | --- |
| **Bone Health** | What are risk factors for low bone density/fracture in adults with CP (e.g., sex, age, diet/ nutrition, mobility function, medications, especially antiseizure medications)?<br>How do bone health and fracture history relate to fall history, fall risk, and fear of falling? How do *near* falls impact bone health and fracture risk?<br>Do prior falls result in behavior change (e.g., walk less to avoid falling, other avoidance behaviors)? How does one's level of risk avoidance/risk tolerance impact their bone health and fracture risk? | Would preventive measures in childhood, such as weight training and ensuring adequate nutrition, mitigate fracture risk in adulthood?<br>What is the appropriate prevention protocol to minimize fracture risk in adults with CP (e.g., exercise, nutrition, supplements)?<br>What is the appropriate treatment threshold and target level for vitamin D in adults with CP?<br>How does access to equipment impact fracture risk? (e.g., Do standers, orthotics, lifts reducing fracture risk?) | What are specific recommendations/ guidelines for bone density evaluation in adults with CP?<br>How can we incorporate screening for bone health risk factors into routine clinical practice? (e.g., calcium and vitamin D intake, weightbearing, fracture history, nutritional status)?<br>When should adults with CP get a bone density scan? How often? Will this change management of their bone health? | What is the appropriate treatment protocol for individuals that have low bone density or have had fractures (e.g., therapy protocol, exercise, nutrition, supplements, medications)?<br>What is the treatment threshold? Does this differ based on an individual's characteristics or comorbidities?<br>How does *recovery* from fracture differ for adults with CP compared to adults without CP? Does this change with age? With bone density? |
| **Kidney Health** | What factors affect creatinine level in adults with CP (e.g., medications, supplements, exercise)?<br>What are the risk factors for kidney disease in adults with CP (e.g., medications, diet, social determinants of health, renal stones, bladder issues)? | What impact does chronic under hydration have on renal function? How do societal factors contribute to this (e.g., less hydration → less urine output → less need for an accessible restroom or for changes by a caregiver)? | Do adults with CP need a different "normal" creatinine range, or should changes over time be monitored?<br>Can we "correct" creatinine for amount of muscle mass? How would this vary based on an individual's GMFCS level? |  |
|  | Particularly for women, how do kidney health, urologic health, and reproductive health relate?<br>How does mobility level affect bladder and bowel function, and in turn kidney health? |  | What percentage of change in creatinine should prompt referral to a nephrologist?<br>Can ultrasound be a useful screening tool to evaluate for residual urine, asymptomatic renal stones? |  |
| **Preventive Care** | How can we improve medical education and training to minimize ableism in the healthcare setting?<br>How does geography and rurality of one's location impact access to preventive care?<br>Does the prevalence or incidence of cancer differ among adults with CP compared to adults without CP?<br>Does the prevalence or incidence of drug use differ among adults with CP compared to adults without CP? | How can we empower and educate community members to be advocates in their care, when they do see a provider that is unfamiliar with CP?<br>How can we support PCPs in busy clinics, seeing patients with a huge range of diagnoses, in caring for the unique needs of adults with CP? | What are appropriate preventive screening guidelines, specific to adults with CP? How can we stratify these based on individual characteristics and risk factors? | How can we leverage existing care teams to include providers knowledgeable about CP, even if the PCP does not have this specialized expertise?<br>Does access to providers who are cross trained between pediatric and adult specialties improve the care of adults with CP? |

GMFCS – Gross Motor Function Classification System

### Bone Health

Many participants with lived experience had personal experience with bone health issues, and many clinicians found these issues extremely relevant to their practice. Community leaders also felt that these issues were important to both themselves and members of their communities. Despite experiencing multiple fractures, fractures from minor trauma, and low bone mineral density, those with lived experience also shared difficulty finding providers knowledgeable about and comfortable with their needs.

Since providers were unaware of the bone health risks facing adults with CP, they were unable to provide anticipatory guidance or lead screening, evaluation, or management interventions to detect and/or treat bone health issues. Clinician participants expressed uncertainty regarding best practices for bone health screening — for example, when to get a bone density scan, how often, and if/when to initiate medications for bone health. Furthermore, adults with CP shared that they, too, were unaware of these risks until they had already experienced a fracture. For example, one participant broke her shoulder, requiring her to be in the hospital for three weeks and then in a long-term care facility for a month. Prior to this accident, bone health had never been discussed.

### Kidney Health

The increased risk of chronic kidney disease was new information for many participants with lived experience, and they consequently felt that increasing awareness among the community as well as providers was critical. A couple of adults with CP reported personal experience with renal issues, including nephrolithiasis and congenital kidney disease. Several clinical investigators shared that they saw issues like nephrolithiasis and bladder issues (e.g., urinary retention, chronic infections) more frequently than frank renal disease.

Interestingly, several participants brought up societal and environmental factors that may influence kidney health. The lack of accessible restrooms and/or assistance to use the restroom may cause some adults with CP to withhold from urinating, or may even incentivize some to purposefully underhydrate themselves to avoid needing to urinate in public. One of these participants recalled an experience where she did not go the bathroom for a whole day in college because of the lack of accessible bathrooms. Several clinical investigators also discussed the impact of chronic underhydration on renal health in adults with CP, and similarly tied in societal factors.

### Preventive Care

Many participants across all focus groups felt that the issues and barriers surrounding preventive care resonated with their experiences. Multiple participants in the lived experience and community leader groups felt that the transition period between pediatrics and adulthood was a particularly challenging time given changes in providers and services, and the inaccurate view of CP as a static, childhood condition. Participants again highlighted lack of provider awareness as a barrier to comprehensive care and emphasized the importance of educating healthcare professionals as well as the CP community about the unique health needs and risks of adults with CP. Community awareness can help facilitate advocacy with providers who may be unfamiliar with CP.

Several participants highlighted the significant accessibility barriers to preventive care, particularly when it comes to women’s health (e.g., mammograms and pelvic exams). In this case, even if the PCP orders the appropriate screening test, if the healthcare facility itself is inaccessible, then the test cannot be completed. Participants noted other barriers to preventive care, including the time pressure of short preventive care visits, insurance barriers, access to a primary care provider (PCP). Clinical investigators also found these barriers and challenges relevant to their clinical practice, and noted particular challenges for adults with CP in accessing dental care, nutritional counseling, and sexual and reproductive healthcare.

## Discussion

We sought to explore stakeholder experiences and identify research priorities for the adult cerebral palsy (CP) research agenda to co-create a multicenter research program focused on adults and aging with CP. Using the infrastructure of the Cerebral Palsy Research Network (CPRN), we conducted iterative focus groups with adults with CP, caregivers, clinical investigators, and community leaders using a qualitative descriptive approach. Participants identified issues related to preventive care as the top priority, though also found bone and kidney health to be important issues. Through thematic analysis of data, we identified themes that cut across these various research areas related to patient and provider knowledge gaps, a precision medicine approach for adult CP care, and the need to address ableism.

Adults with CP face unique health risks, including greater risk of stroke and myelopathy; high rates of chronic fatigue and chronic pain; and high prevalence of many cardiovascular, metabolic, respiratory, musculoskeletal, and psychological chronic conditions, as well as multimorbidity.^2–8^ Additionally, compared to adults without CP, adults with CP have higher healthcare utilization across all settings and 154% higher all-cause healthcare costs.^41^ However, despite this high burden of chronic disease and healthcare utilization, the vast majority of clinical services, interventions, funding, and research studies are focused on children, and CP in adulthood has been the focus of only 4% of all CP-related National Institutes of Health funding.^42–44^ In 2018, the CPRN set a patient-centered research agenda for CP, including consumers, clinicians, and researchers in the process, and identified issues related to aging with CP as the top research priority.^45^ Our study built upon and expanded this top priority by exploring specific topic areas related to aging with CP (bone health, kidney health, and preventive care) and is the first to engage the adult CP community in doing so. Preventive care emerged as the top priority among those discussed in our study, and improvements in preventive care can facilitate the earlier identification and treatment of many chronic diseases.

Participants throughout all focus groups in our study cited a lack of provider – and patient – knowledge as a significant barrier to appropriate care. Many participants with lived experience were learning of these unique health risks for the first time during our study, and many also recalled having to educate their providers about CP. Prior studies have also found a lack of knowledgeable providers to be a barrier to adult CP care.^10,11^ In addition to lack of disability-specific knowledge, multiple studies have also found a lack of adult provider *comfort* with developmental disabilities to be a major barrier to age-appropriate care.^46–48^ Our participants supported these findings, recounting clinical encounters with providers who did not feel comfortable with their needs but also with providers who were willing to engage and support them despite a lack of CP “expertise”. While there have been many educational programs designed to improve training of medical and health professional students in caring for persons with disabilities, and intellectual and developmental disabilities (IDD), in particular, many barriers to instituting mandated disability education and training requirements persist (e.g., overfull curriculums, inadequate resources, lack of faculty champions).^47,49–52^ One study that audited the IDD content of Australian universities’ medical training found particularly low presence of curricula on preventive health, sexual health, and women’s health, which reflected the care gaps noted by our participants. Interestingly, participants in our study, particularly adults with CP, also emphasized the need to educate patients themselves on the unique risks and needs facing adults with CP. Many adults with CP shared that they are often their own best advocate in their care and desire CP-specific knowledge to continue to advocate with their care teams. Empowering patients to self-advocate can improve communication and enhance patient-centered care, goal-concordant care, and both patient and provider experience.^53,54^ Therefore, educational efforts should be bidirectional, with efforts geared towards both health professionals and persons with lived experience.

CP is an extremely heterogeneous condition, and individuals with CP can have multiple types and severities of functional impairments and medical comorbidities.^55–58^ Given this heterogeneity, participants in our study highlighted the importance of a precision medicine approach to care for adults with CP, taking into account an individual’s risk factors when making preventive, screening, and treatment recommendations. While precision medicine is often used in reference to genomics and biomarkers, its definition more broadly speaks to “individualizing the health-care process to the uniquely evolving health status of each patient.”^59^ And while the use of genomics and/or biomarkers to guide CP care may be a long-term future goal, participants spoke to consideration of variables like functional level, age, sex, medical history, and medication use in tailoring an individual’s care, thereby providing more effective and efficient care. In disability healthcare and rehabilitation, precision rehabilitation focuses on delivering the “right intervention, at the right time, to the right individual” to maintain and optimize function.^60^ Similar frameworks have been described to tailor rehabilitation interventions to the individual in spinal cord injury and traumatic brain injury and to guide physical therapy interventions.^61–63^ An example discussed by our participants focused on personalized approaches to bone density screening (e.g., using age, sex, mobility status, fracture history, fall risk to guide initiation and frequency of screening). However, such an individualized screening pathway would require large sample sizes and likely long-term follow up. Near-term future studies should focus on exploring likely high-yield risk factors to guide care, with the long-term goal of a more detailed, personalized approach.

Ableism influences many of the complex, multifactorial barriers to equitable healthcare that people with disabilities, including CP, face.^64–67^ Indeed, adults with CP have lower rates of annual wellness visits; experience fractured, inconsistent care; and have healthcare teams largely driven by availability of providers rather than expertise or comfort.^9,10,24,25,68^ Prior studies have shown that most physicians believe that people with significant disability have worse quality of life than those without disabilities; less than half of physicians were very confident in their ability to provide equal care to persons with disabilities; and physicians often feel overwhelmed by the demands of providing care to people with disabilities.^64,65^ In addition to the lack of provider knowledge discussed above, our participants discussed the lack of adequate time in clinical visits to discuss their needs and concerns adequately, ableist assumptions about sexual and reproductive health, and concerning communication challenges (e.g., fear that clinicians would assume a patient is “drunk” due to speech difficulties). Some of these barriers may be particularly impactful in the primary care setting, where lack of time, poor reimbursement, and care coordination burdens particularly impede access for patients with disabilities.^69^ And while participants acknowledged these factors, several also emphasized the importance of facilitating comprehensive primary care for adults with CP, rather than delegating CP care to specialists, since primary care is frequently the gateway to specialty care.

### Study Limitations

This study has several important limitations. Participants were recruited from CP- and disability-focused networks, and therefore our findings may not be transferable to the general community, specifically to people who are less connected with and less active in CP research and advocacy efforts.^70^ Similarly, while we were able to recruit participants with lived experience who were non-White and Hispanic, the majority of lived experience participants were White and non-Hispanic. While it is important to amplify the voices of adults with CP in this type of research, our lived experience focus groups only included one caregiver; therefore, the perspectives of a larger group of caregivers, who often care for individuals with more severe disability, may differ. We successfully recruited adults with CP with varying GMFCS levels; however, we did not have sufficient numbers of participants within each GMFCS level to directly compare between subgroups. Future research could evaluate whether or not research priorities vary based on one’s functional mobility level.

### Conclusions

Adults with CP face unique healthcare needs and risks as they age, and the evidence base to guide their care lags significantly behind. Individuals with lived experience, clinical investigators, and community leaders in our study identified preventive care as the top research priority for the adult CP research agenda. Knowledge gaps among patients and providers alike, the desire for a precision medicine approach to guide care for adults with CP, and the impact of ableism on care impacted all potential research areas discussed. Next steps in this line of research should focus on interventions to facilitate primary and preventive care interactions for adults with CP while continuing to engage stakeholders in the research process. The research goals and design should directly attend to the insights gained in this study. Participants also felt that bone health was a significant issue, and future research should focus on delineating who is most at risk for fractures and low bone mineral density to guide appropriate screening measures.

## Acknowledgements

This work was supported by the Cerebral Palsy Alliance Research Foundation (CPARF) who funded an Accelerator Award for work within the Cerebral Palsy Research Network. The funder had no role in determining the funding, design, implementation or interpretation of the findings.

We would like to thank the adults with cerebral palsy, caregivers, clinical investigators, and community leaders who participated in our study, sharing their experiences and perspectives with the hopes of improving health, function, and quality of life for individuals with cerebral palsy across the lifespan.

## Data Availability Statement

The data that support the findings of this study are available from the corresponding author upon reasonable request.

